# Characteristics and Preparedness for COVID-19 Outbreaks of Australian Residential Aged Care Facilities: a Cross-sectional Survey

**DOI:** 10.1101/2022.02.05.22270416

**Authors:** Gwendolyn L Gilbert, Jenna Smith, Erin Cvejic, Alan Lilly, Kirsten McCaffery

## Abstract

**Objective:** To provide an overview of Australian residential aged care facilities’ (RACFs’) COVID-19 outbreak preparedness and responses, 12 months after the pandemic began in early 2020.

**Methods:** A cross-sectional survey of RACF managers was conducted as part of an overview of COVID-19 experience during 2020. Survey questions were based on findings of previous outbreak reviews.

**Results:** Comparison with available data from the Australian Institute for Health and Welfare suggested that survey respondents (n=331) were a representative sample. Almost all RACFs had outbreak management plans, including provision for a surge workforce. However, anticipated staff replacements fell short of those often required during outbreaks. Staff of most (83%) RACFs had completed online infection control training, and a smaller proportion (73%) face-to-face training, by the time of the survey. Exploratory analyses to identify RACF characteristics associated with increased outbreak risk found a strong association with location in Victoria (adjusted risk ratio [aRR] 12.8) where most community transmission occurred during 2020. The only other association was an increased risk in facilities where all staff had not completed face-to-face infection control training (aRR 2.1). Respondents ranked leadership and management; planning and preparation; and infection control as the top three of seven critical lines of defence against COVID-19.

**Conclusion:** Survey results suggest that, in early 2021, most Australian RACFs were better prepared for the ongoing risk of COVID-19 than in 2020. Continued implementation of the Aged Care Royal Commission’s recommendations is needed to ensure the aged care sector is prepared for future infectious disease emergencies.

## INTRODUCTION

Based on experience in China and Italy in early 2020,(1, 2) it was apparent that older people, particularly those in residential care, would be most severely affected by COVID-19. The index case of the first outbreak in an Australian residential aged care facility (RACF) was diagnosed on March 3 2020 in a nursing assistant at Dorothy Henderson Lodge, on Sydney’s North Shore.(3) Two residents had been admitted to hospital several days before with serious illnesses that were later diagnosed as COVID-19. The total number of cases in that outbreak was limited to five staff members and 17 residents (of whom six died) by the decisive actions of facility management and public health authorities, including rapid implementation of strict infection prevention and control (IPC) measures and transfer of infected residents to hospital.(4)

Another outbreak, at Newmarch House in outer western Sydney in April 2020, involved 34 staff and 37 residents, of whom 19 died. Most infected residents were managed in the facility, by the hospital-in-the-home team from the local hospital. However, a large proportion of staff, who were close contacts of cases, had to be quarantined and finding suitable replacements was difficult. Many residents who remained uninfected suffered serious adverse effects from isolation, immobility and inadequate care. Delayed implementation of effective IPC measures and the continued presence of infected residents in the home contributed to ongoing spread of infection.(5)

During the first wave of COVID-19 (March-June, 2020) in Australia there were more than 60 RACF outbreaks – defined as one or more cases in a staff member, resident or frequent visitor. Apart from the two major Sydney outbreaks and one in Melbourne in June (45 staff and 37 residents, with 15 deaths), these outbreaks involved fewer than five cases.(6) However, when the second wave began in Victoria, in June 2020, the situation rapidly changed. Of 226 outbreaks during this period, 50 accounted for the majority of infections among residents, ranging from 10 to 102 cases. By mid-September 2020, approximately 2050 residents had been infected and 655 had died. In addition, around 2200 RACF staff had been infected, ranging from eight to 131 across outbreaks, but many more were furloughed and replacement staff were often poorly qualified and inexperienced.

During 2020 and early 2021, the Commonwealth Department of Health (DoH) commissioned two of the authors (GLG and AL) to undertake reviews of four major RACF outbreaks (4, 5, 7) and of COVID-19 outbreaks in Australian RACFs, generally.(8) The aim of these reviews was to identify lessons learnt from failures of outbreak control in a minority of RACFs and key factors that contributed to prevention or rapid control of outbreaks in others. The individual RACF outbreak reviews (5, 7), identified seven *lines of defence* that were critical to successful outbreak control but lacking, to varying degrees, in those where major COVID-19 outbreaks occurred. These critical defences were: leadership and management; effective communication; planning and preparation; infection control; pathology testing; workforce; emergency management.

## AIMS AND OBJECTIVES

This online survey was part of the national review of COVID-19 outbreaks in RACFs. Its aim was to provide a broad overview of the preparedness for, and experience of COVID-19 outbreaks in RACFs from the perspective of senior facility managers. It complemented the findings of interviews and workshops with key stakeholders that contributed to the review.

## METHODS

### Particpants and recruitment

Particpants were senior managers or directors of nursing of Australian RACFs. The survey was initially advertised in the DoH’s newsletter to aged care providers. An invitation to participate, with a link to the online survey, was emailed on February 23, with a reminder on March 5 2021, to the DoH Bulk Information Distribution Systems (BIDS) mailing list, to which representatives (managers/senior staff) of most individual RACFs subscribe. The survey was open from March 2 to 17 2021. Survey questions were accessible only to recipients who agreed to participate.

### Survey design

The study was a cross-sectional survey administered using Qualitrics survey software. The research team and a convenience sample of volunteers pilot-tested the instrument to assess the time taken to complete it and the clarity and sequence of questions. The survey was designed to be completed in 10-15 minutes. The landing page of the survey included a participant information sheet.

### Measures

Survey questions were based on findings of previous reviews by GLG and AL (4, 5, 7) and refined in collaboration with author KMcC. The survey instrument contained questions relating to characteristics of respondents and the RACFs they represented and aspects of the facilities’ preparedness for future COVID-19 outbreaks. Respondents who indicated that their facility had experienced a COVID-19 outbreak during 2020 were asked a short set of additional questions. The full survey instrument is available in Supplementary Materials.

### Statistical analyses

Descriptive analyses (frequency and relative frequency; medians and interquartile ranges [IQR]) were used to examine characteristics of respondents and the facilities they represented and outcomes of COVID-19 outbreaks.

Exploratory analyses were conducted to identify potential organisational factors associated with RACFs that experienced at least one COVID-19 case. Nine exploratory variables were included in a generalised linear model (modified Poisson approach, with a log-link function and robust standard errors) to estimate relative risks and corresponding 95% confidence intervals. Sub-group analyses were conducted for RACFs located in Victoria only, and repeated using a more restricted definition of an outbreak (i.e. five or more resident and/or staff cases).

Statistical analyses were conducted using Stata/IC v16.1 (StataCorp, College Station, TX, USA). A p-value of less than (<) 0.05 was set as the threshold for statistical significance.

### Ethics

This survey was approved by the Commonwealth DoH as part of the review of COVID-19 outbreaks in Australian RACFs. The review was widely publicised in the aged care sector and potential participants were assured that any information provided would contribute to the findings of our review but not attributable to any individual without express permission. By completing and returning the survey, participants consented to these conditions. The full review report was published on the DOH website and on November 1 2021 and is publicly available.(8)

The survey was referred to only briefly in the review report (8) but detailed findings were not included. This report fulfils our commitment to survey participants that the findings would be made available to them and the aged care sector, generally.

The survey was a quality assurance and evaluation project which complied with criteria outlined in the NHMRC Ethical Considerations in Quality Assurance and Evaluation Activities (section 2, p2-4).(9). Data collected were coincidental to RACF standard operating procedures (routine organisational information and COVID-19 response situational factors). Only self-reported data were analysed, for the purpose of identifying areas for improvement in RACF outbreak preparedness, and not linked to any specific individual or organisation; the privacy and professional reputations of providers were maintained. (9)p.4 DoH officials had no part in the design, analysis or reporting of the survey, but agreed to publication of the results.

The University of Sydney Human Research Ethics Committee approved the project, retrospectively, on January 13, 2022 (project no. 2021/799).

## RESULTS

### Responses

Survey responses were included in the analysis if a) at least 75% of the survey questions were completed; b) the RACFs represented had more than five registered places; and c) there were no logical inconsistencies indicative of potentially unreliable responses (e.g. the number of registered places in the facility exceeded 400 or the number of reported resident cases or deaths exceeded the number of registered places in the facility). Of 685 responses received, 354 were excluded, leaving 331 valid responses for analysis.

We could not determine accurately how many of the 2,722 RACFs in Australia, as of 30 June 2019,(10) were represented among subscribers to the DoH BIDS. Assuming that the number of surveys distributed to eligible respondents was approximately 2500, the rate of valid responses was ∼13%.

The jurisdictional distribution of valid responses was: New South Wales 108 (33%); Victoria 92 (28%); Queensland 59 (18%); Western Australia 33 (10%); South Australia 24 (7%); Tasmania 6 (2%); Australian Capital Territory 6 (2%); Northern Territory 3 (1%).

### Characteristics of respondents and RACFs represented

Characteristics of respondents (N=331) included in the analysis, and the RACFs they represented, are shown in Table 1. Most respondents (80%) were facility managers and/or directors of nursing and 74% held a nursing qualification; the RACFs they represented had between six and 330 registered places. The median number of registered places, their State/Territory locations and the proportions of RACFs with various types of funding arrangements were consistent with those recorded by the Australian Institute of Health and Welfare (AIHW) GEN Aged Care Data (2020) - *Services and Places in Aged Care, 30 June 2019*.(10)

**Table 1.**
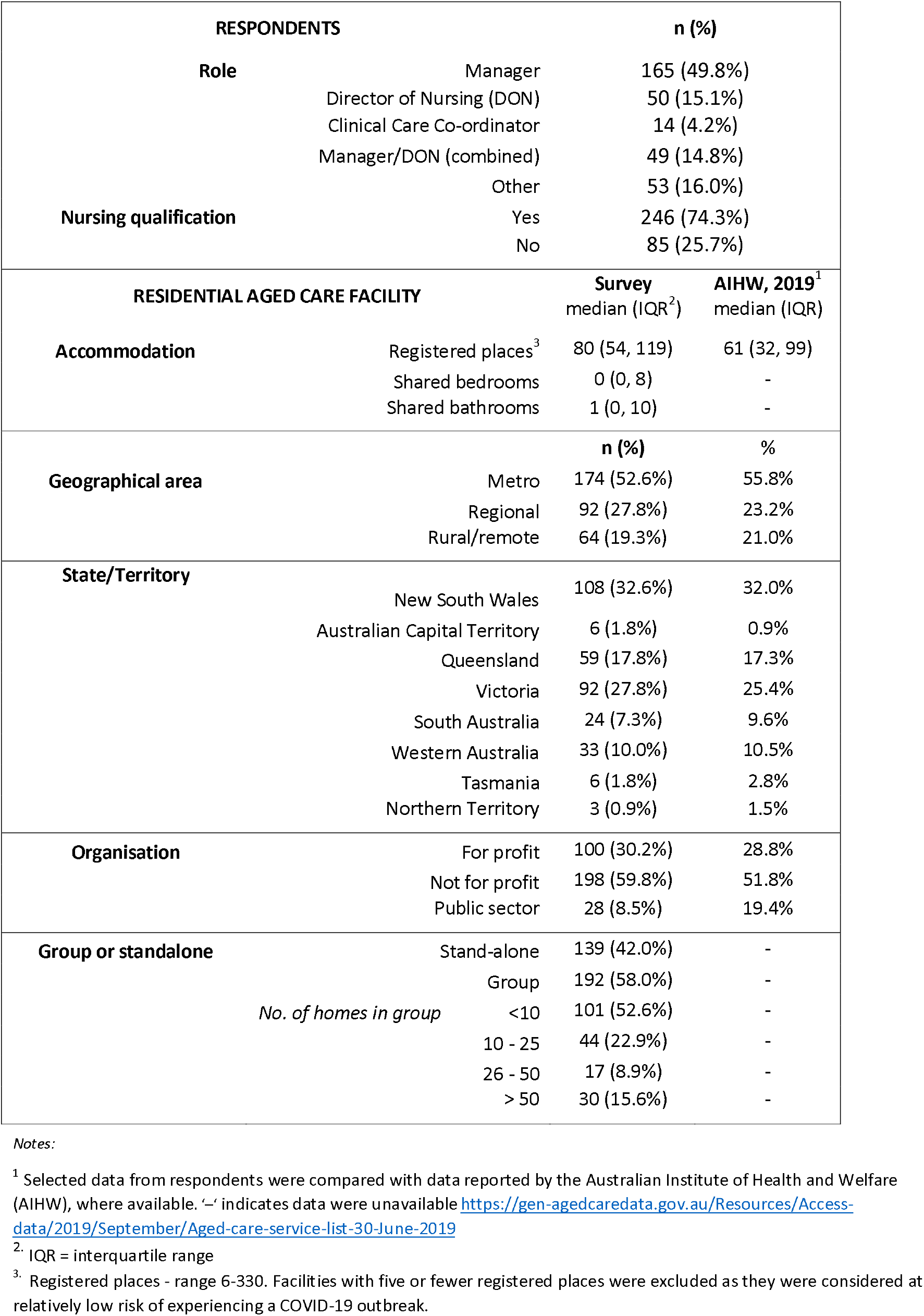
Characteristics of survey respondents and the residential aged care facilities (RACFs) they represented (N=331).

Table 2 summarises responses to questions relating to A) information provided by official sources; B) IPC training and C) OMPs including D) provision for a surge workforce.

**Table 2.** Survey respondents’ answers to selected survey questions (number of responses)

|  |  |  |
| --- | --- | --- |
| **Q. A) How satisfied were you with information provided and delivery?** |  | **(n=331)** |
| **Source of information** |  |  |
| *Commonwealth Department of Health* |  |  |
|  | Very/quite dissatisfied | 71 (21.5%) |
|  | Neutral | 62 (18.7 %) |
|  | Very/quite satisfied | 197 (59.5%) |
|  | Unaware of advice | 1 (0.3%) |
| *State/Territory Department of Health* |  |  |
|  | Very /quite dissatisfied | 52 (15.7%) |
|  | Neutral | 52 (15.7%) |
|  | Very/quite satisfied | 222 (67.1%) |
|  | Unaware of advice | 5 (1.5%) |
| *State/Territory Public Health Unit* |  |  |
|  | Very/quite dissatisfied | 45 (13.6%) |
|  | Neutral | 54 (16.3 %) |
|  | Very/quite satisfied | 218 (65.9%) |
|  | Unaware of advice | 14 (4.2%) |
| *Aged Care Quality and Safety Commission* |  |  |
|  | Very/quite dissatisfied | 87 (26.3%) |
|  | Neutral | 72 (21.8%) |
|  | Very /quite satisfied | 169 (51.1%) |
|  | Unaware of advice | 3 (0.9%) |
| **Q. B) What proportion of your staff completed:** |  | **(n=331)** |
| *Online IPC training?* |  |  |
|  | All (>90%) | 276 (83.4%) |
|  | Most (61-90%) | 40 (12.1%) |
|  | Some (40-60%) | 11 (3.3%) |
|  | Few (<40%) | 4 (1.2%) |
| *Face-to-face training?* |  |  |
|  | All (> 90%) | 242 (73.1%) |
|  | Most (61-90%) | 62 (18.7%) |
|  | Some (40-60%) | 16 (4.8%) |
|  | Few (<40%) | 11 (3.3%) |
| **Q. C) For RACFs with an OMP (99% of total)** |  | **(n=329)** |
| *OMP developed before July 31 2020?* |  | 266 (80.9%) |
| *OMP practiced during 2020?* |  | 239 (72.6%) |
| **Q. D) For RACFs with surge workforce plan (91% of total)** |  | **(n=300)** |
| *Anticipated percentage of staff replacements?* |  |  |
| $\leq 30\%$: | Before July 31 2020 | 147 (49.0%) |
|  | After July 31 2020 | 98 (32.7%) |
| 30-50% | Before July 31 2020 | 98 (32.7%) |
|  | After July 31 2020 | 117 (39.0%) |
| >50% | Before July 31 2020 | 55 (18.3%) |
|  | After July 31 2020 | 85 (28.3%) |
| *Did surge staff receive special training?* |  | 146 (48.7%) |

The highest levels of satisfaction were with infomation provided by State/Territory Health Departments and Public Health units, and the lowest with that from the Aged Care Quality and Safety Commission (ACQSC).

Most (83%) respondents reported that virtually all staff (>90%) in their facilities had completed online IPC training. Face-to-face training had been completed by all staff in a smaller proportion (73%) of facilities. The survey did not ask when IPC training had occurred.

All but two (n=329) respondents indicated that their facilties had OMPs, of which 81% had been developed before July 31 2020, 73% had been practiced at least once and, of those, 95% had modified them as a result. There was a high level of confidence that OMPs were fit for purpose. 300 (91%) facilities had contingency plans for an internal surge workforce, but the anticipated proportion of staff replacements changed over time. Before July 31 2020, 18% of respondents had plans to replace 50% or more staff; this proportion increased to 28% after July 31 2020. Fewer than half of the respondents indicated that internal surge workers had received specific training.

### What counts most in defence against COVID-19 outbreaks?

Respondents were asked to rank the seven critical ‘lines of defence’, identified in previous reviews, in order of importance from one (most) to seven (least). The results are shown in Figure 1. Defences ranked most highly by the greatest proportions of respondents were leadership and management (32%), planning and preparation (28%) and infection control (25%).

**Figure 1.**
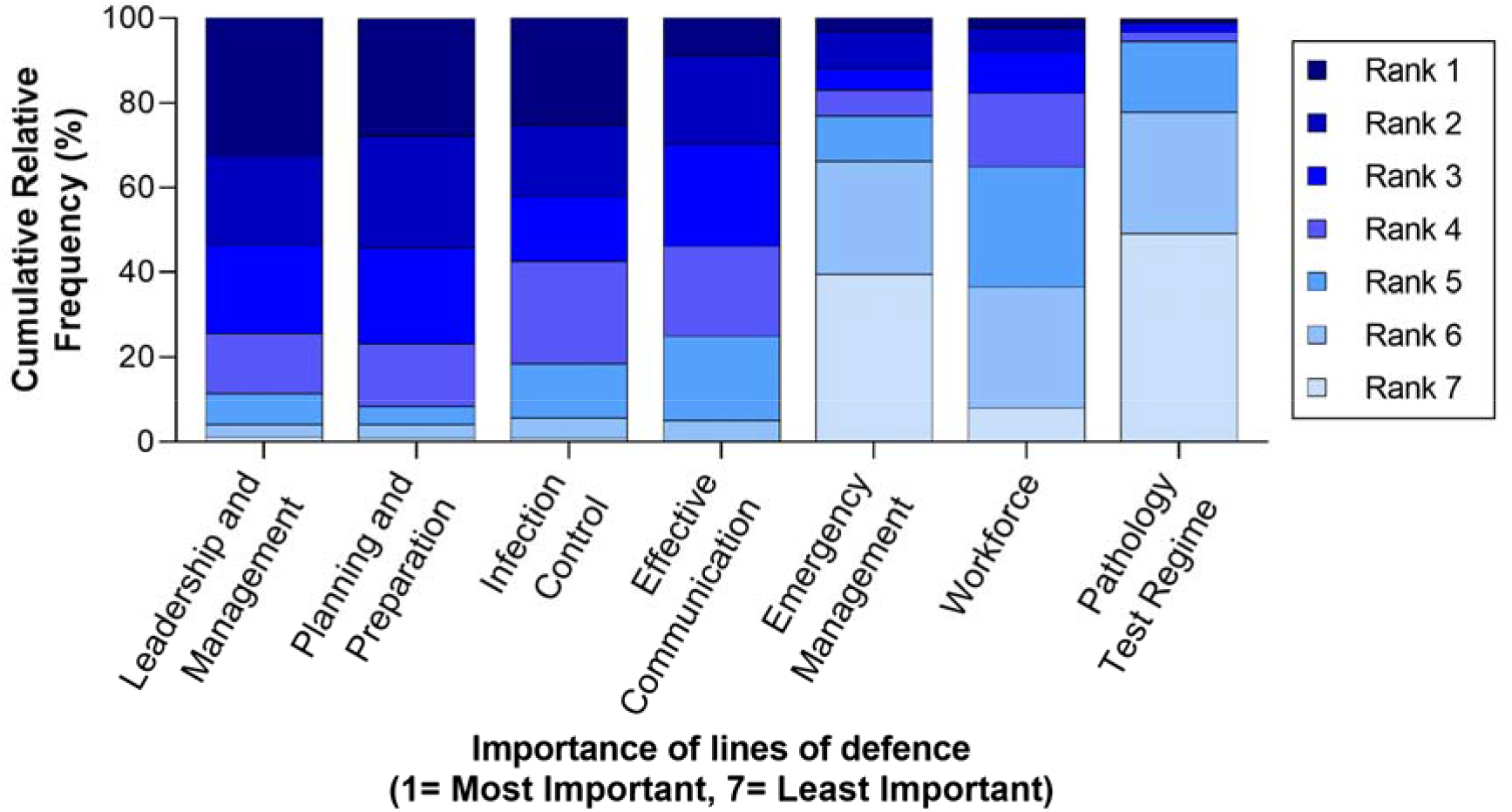
Cumulative relative frequency of perceived importance of different lines of defence, ranked from 1 (most important) to 7 (least important). “Leadership and management” was most frequently ranked as most important (32.3%), followed by “planning and preparation” (27.9%) and “infection control” (25.1%).

### COVID-19 outbreaks

Of 331 facilities represented in the survey, 32 (10%) had experienced outbreaks, of which 19 (6%) involved five or more cases (suggesting SARS-CoV-2 transmission within the RACF). Twenty seven (84%) outbreaks were in Victoria, three (9%) in New South Wales and one each in Queensland and Tasmania. Some outcomes and effects of these outbreaks are summarised in Table 3.

**Table 3.** Outcomes and effects in residential aged care facilities that experienced at least one COVID-19 outbreak (n=32) ^*^.

| **OUTCOME** | **median (range; IQR)** |
| --- | --- |
| Resident cases | 11 (0-90; 1,30) |
| Resident deaths | 3 (0-38; 0,11.5) |
| Staff cases | 10.5 (0-53; 2,40) |
| **EFFECT** | **n (%)** |
| **Q. What proportion of time could you maintain an adequate workforce during the outbreak?** |  |
| >90% | 12 (37.5%) |
| 61-90% | 8 (25.0%) |
| 40-60% | 9 (28.1%) |
| <40% | 3 (9.4%) |
| **Q. Did any staff stay away from work because of concern for their health/wellbeing?** |  |
| <20% | 19 (59%) |
| >20% | 13 (41%) |
| **Q. How much of the time did you have timely access to adequate PPE supplies?** |  |
| > 90% | 16 (50%) |
| 61-90% | 7 (22%) |
| 40-60% | 7 (22%) |
| < 40% | 2 (6%) |
| **Q. How much of the time were you able to maintain adequate documentation of resident care?** |  |
| >90% | 12 (38%) |
| 61-90% | 11 (34%) |
| 40-60% | 4 (13%) |
| <40% | 5 (16%) |
| **Q. What type of resident care records system did you use?** |  |
| Electronic | 17 (53%) |
| Paper-based | 4 (13%) |
| Some of both | 11 (34%) |
| **Q. Did residents wear any form of identification during outbreak?** |  |
| No | 19 (59%) |

\*32 of 331 (9.7%) RACFs experienced an outbreak based on the official definition (one or more COVID-19 cases)

There were wide variations but, overall, resident and staff case numbers were similar and there was a positive correlation between them, within individual RACFs (Spearman’s *ρ*= 0.82,p< 0.001). There was also wide variation in the numbers of resident deaths.

Most facilities were able to maintain an adequate workforce for at least 60% of the time during the outbreak, but a significant minority could not. Some staff absenteeism was due to concern for their health and well-being. Timely access to adequate PPE was problematic for some RACFs.

Electronic resident care records were used, with or without additional paper-based records, by most facilities; documentation of resident care was maintained for most of the time during outbreaks. A majority of residents (59%) did not wear any form of identification, such as a wristband.

*Additional outbreak findings*: 26 (81%) RACFs engaged extra staff to reduce residents’ loneliness during outbreaks; 24 (75%) monitored residents’ daily food and fluid intake; 22 (69%) undertook structured outbreak reviews and, of these, 18 (82%) provided a report to the organisational executive and 16 (73%) to the Board.

### Exploratory analyses of organisational factors associated with COVID-19 outbreaks

Exploratory analyses were conducted for 328 facilities for which data on all nine of the following variables were available: State/Territory (Victoria/ other); location (non-metropolitan/ metropolitan); organisational ownership (for profit/ not-for-profit); organisational structure (standalone/ group); OMP developed (none/ late/ early); OMP tested (none or not tested/ tested); registered places (≥75/ <75); shared rooms or bathrooms (both/ either/ none); staff completed online training (<90%/ >90%); staff completed face-to-face training (<90%/ >90%). Results are shown in Supplementary Table 1.

RACFs in Victoria were at much higher risk of outbreaks (adjusted relative risk [aRR] 2.08; 95% confidence interval [CI] 4.74-30.75; p<.001). After adjusting for all other variables, the only other organisational factor associated with an increased outbreak risk was less than 90% of staff having completed IPC training (aRR: 2.10; 95%CI: 1.17-3.79; p=.013). The results were similar for a sub-group analysis of the 91 RACFs located in Victoria that provided complete data, of which 27 (30%) experienced outbreaks. Again, the only significant association with an outbreak was less than 90% of staff having completed face-to-face IPC training (aRR: 2.20; 95%CI: 1.15-4.19; p=.017).

The exploratory analysis was repeated using a more restricted COVID-19 outbreak definition, namely five or more resident and/or staff cases. The results are shown in Supplementary Table 2. They should be interpreted with caution since the outbreak definition was arbitrary, the number of larger outbreaks (the outcome of interest) was small (n=19) and the timing of outbreaks relative to staff training unknown. In this subgroup the risk was higher for Victoria RACFs (aRR: 35.95; 95%CI: 4.25, 304.11; p=.001), but less for facilities outside, than for those within, metropolitan areas (aRR: 0.05; 95%CI: 0.01, 0.32; p=.002). The association between outbreaks and <90% of staff having completed face-to-face training failed to reach statistical significance (aRR: 2.58, 95%CI: 1.00, 6.67; p=.051).

## DISCUSSION

This survey provided useful insights into some characteristics of Australian RACFs relevant to their experience with COVID-19 in 2020. Based on comparison with the most recent data available from the AIHW,(10) the RACFs surveyed were a representative sample, with respect to jurisdictional distribution; metropolitan/non-metropolitan location; number of registered places; and organisational type.

From early 2020, when the SARS-CoV-2 pandemic began, RACFs were identified as potentially high risk settings and aged care providers were provided with information and guidelines about outbreak prevention, planning and management from various sources, including Commonwealth and State health departments and the ACQSC. (11) As international and local experience with RACF outbreaks increased, (4, 12-14) advice was modified. However, the frequency of change and inconsistencies between sources caused confusion and frustration among RACF managers.(5, 7) This was confirmed by survey respondents, some of whom reported dissatisfaction with the quality and/or delivery of information. Respondents were most satisfied with material from State Health Departments and/or public health units, which may reflect more specific, up-to-date knowledge of local RACF conditions.

Interim international and local IPC guidelines for RACFs were also issued early in 2020 and updated periodically.(15-17) Online training was provided by Commonwealth and most State authorities and RACF managers were urged to ensure that all staff completed it. An online self-assessment survey by the ACQSC in March 2020, found that virtually all aged care providers believed their organisations’ IPC preparations were appropriate. However, the Royal Commission into Aged Care (RCAC) Quality and Safety’s special report ‘Aged Care and COVID-19’ in October 2020, noted that many RACFs lacked adequate IPC expertise, a culture of compliance and basic IPC education and training.(18) Even providers who recognised the importance of IPC, admitted to having under-estimated the enormous impact of COVID-19, including the quantities of PPE that had to be purchased, stored and disposed of after use, and the increased workload and discomfort for staff using it.(19) By contrast, aged care sectors in countries that had experienced outbreaks of SARS (2003) and/or MERS (2015), including Hong Kong, Singapore, Taiwan and South Korea, had developed robust IPC measures before the pandemic began, and successfully limited the burden of COVID-19.(12-14, 20-23)

The majority of respondents reported that virtually all staff (>90%) in their facilities had completed online (83% of respondents) and/or face-to-face (73% of respondents) training. However, the survey did not ask *when* training had occurred. Based on reports from participants in previous reviews,(5, 7) and the national review,(8) staff of many RACFs had had limited, if any, COVID-19-specific IPC training by mid-2020; most training occurred during or after the Victorian second wave. Based on recommendations by the RCAC, measures have been introduced nationally, to improve IPC training and practice; all RACFs are now required to appoint a nurse, who has completed an approved IPC course, as IPC lead. However, details of their responsibilities, ongoing support and further training are at the discretion of providers.

Most RACFs had an OMP which they had tested and modified before July 31 2020. The survey did not ask for details but most respondents reported being confident that their OMP was fit for purpose. OMPs generally included plans to deploy an internal surge workforce, if required, but only a minority of respondents anticipated a need to replace 50% or more of their staff during an outbreak. The RCAC, in their final report,(24) observed - as had others, previously (25) - that many Australian RACFs were chronically understaffed, even for business-as-usual. When COVID-19 outbreaks occurred many providers had to rely on external resources to replace furloughed staff, and a significant proportion of respondents reported being unable to maintain an adequate workforce.

Local community transmission is the strongest predictor of COVID-19 being introduced into a RACF, often by an asymptomatically-infected staff member.(26) It was therefore not surprising that the majority of outbreaks reported by survey respondents were in Victoria where most community transmission occurred in 2020. In the subgroup analysis of Victorian RACFs, outbreak risk was lower in non-metropolitan than metropolitan facilities. The only other significant finding of the exploratory analyses, was an increased outbreak risk in facilities where not all staff had completed face-to-face IPC training. This may reflect pre-existing IPC standards and staffing ratios (especially registered nurse hours), since *de novo* face-to-face IPC training in the context of a pandemic is particularly challenging.

Large surveys of nursing homes and long-term care facilities (LTCFs) overseas have found that the severity of COVID-19 outbreaks correlates with, among other things, facility size, registered nurse hours, shared accomodation, implementation of recommended IPC precautions and organisational funding.(27-30) Our exploratory analyses did not identify assocations of outbreaks with facility size, organisational funding or shared rooms. However, this probably reflects the relatively small survey size and uneven outbreak distribution, since a study of all 774 nursing homes in Victoria, in the latter half of 2020, found that larger Victorian outbreaks were associated with metropolitan location, larger facility size, shared rooms, and private ownership.(31)

## CONCLUSION

Of the seven critical RACF lines of defence identified in previous reviews, respondents ranked leadership and management, planning and preparation and infection control most highly. Experience with COVID-19 outbreaks in aged care suggests that these are the most important factors that determine an organisation’s resilience. Our findings suggest that, by the time this survey was conducted in early 2021, most Australian RACF managers (and, by inference, aged care providers generally) had recognised the importance of these factors and were better prepared for possible COVID-19 outbreaks. However, there is an ongoing challenge to ensure that the longstanding deficiencies of quality and safety in aged care, identified by the RCAC and further exposed by COVID-19, continue to be addressed and improvements sustained into the future.

## STRENGTHS AND LIMITATIONS

This survey achieved its goal of providing a wide perspective of the preparedness and responses of RACFs across States and Territories and metropolitan, regional and remote areas. Responses to some questions raised additional issues that were covered in more detail by in-depth interviews during RACF outbreak reviews, with smaller samples of aged-care stakeholders.

The survey was limited in scope and detail as we were aware of the enormous stress that participants had endured during the previous year, and from which they were still recovering when the survey was conducted. The initial response from an estimated ∼2500 survey invitation recipients was pleasing. We surmise that the significant proportion of respondents did not complete the survey reflects other, more pressing, commitments.

### Practice/Policy Impact Statement

When the COVID-19 pandemic began many Australian RACFs were ill-prepared. This survey suggests that lessons learnt from serious COVID-19 outbreaks in 2020 contributed to aged care providers’ and government agencies’ being much better prepared in 2021. However, further and sustained improvements in staffing levels, outbreak preparedness and infection prevention and control practices and other measures are required.

## Data Availability

Data are available upon reasonable request. Deidentified participant data will be made available upon request to anyone wishing to access it who provides a methodologically sound proposal to the corresponding author.

## Acknowledgements

The authors wish to thank the Residential Aged Care Managers who responded to this survey.

