## Supplementary materials for "Characteristics and Preparedness for COVID-19 Outbreaks of Australian Residential Aged Care Facilities: a Cross-sectional Survey"

**Survey instrument.**

Introduction

Thank you for your participating in this survey, as part of the national review into COVID-19 outbreaks in residential care.

The survey should be completed by the most senior person at the RACF (most likely Manager or Director of Nursing) with knowledge of local COVID-19 outbreak management plans. If necessary feel free to consult colleagues, but please return only one completed survey per facility.

The data will be collected and analysed by a XXXX researcher. Responses are anonymous and data will be aggregated.

Again, thank you for completing the survey. It is estimated that it will take approximately 10-15 minutes. Your input is critical to continuous improvement in the management of potential COVID-19 outbreaks in residential care.

Respondent characteristics

Which of the following titles best describes your role?

- Manager (1)
- Director of Nursing (2)
- Clinical Care Co-ordinator (3)
- Manager/Director of Nursing (combined role) (4)
- Other (please specify) (5) ______________________________________________

Do you have a nursing qualification?

- Yes (1)
- No (2)

Display This Question:

If Do you have a nursing qualification? = Yes

Are you a registered nurse or enrolled nurse?

- Registered (1)
- Enrolled (2)

RACF characteristics

What is the number of registered places in the RACF you manage? _____________________________________________________________

How many shared bedrooms are there? _______________________

How many shared bathrooms are there? ________________________

Which geographical area best describes the RACF location?

- Metro (1)
- Regional (2)
- Rural (3)
- Remote (4)
- Don't know (5)

In which State or Territory is the RACF?

- NSW (1)
- ACT (2)
- QLD (3)
- VIC (4)
- SA (5)
- WA (6)
- TAS (7)
- NT (8)

What is the postcode of the RACF? (optional) __________________

How would you best describe the facility's organisation?

- For profit (private) (1)
- Not for profit (church or charitable) (2)
- Public sector (government owned) (3)
- Other (please specify) (4) ______________________________

Is the RACF part of a group or a stand-alone facility?

- Group (1)
- Stand-alone (2)

Display This Question:

If Is the RACF part of a group or a stand-alone facility? = Group

Does the group have:

- < 10 homes (1)
- 10 - 25 homes (2)
- 26 - 50 homes (3)
- > 50 homes (4)

Display This Question:

If Is the RACF part of a group or a stand-alone facility? = Group

In which States/Territories are the homes? (Please tick all that apply)

- NSW (1)
- ACT (2)
- QLD (3)
- VIC (4)
- SA (5)
- WA (6)
- TAS (7)
- NT (8)

COVID-19 outbreak management plan (OMP)

Does your RACF have a **COVID-19 outbreak management plan?**

- Yes (1)
- No (2)
- Don't know (3)

COVID-19 OMP questions

When was it developed?

- Before 31 July 2020 (1)
- After 31 July 2020 (2)

Was the plan tested in practice (like you would practice a fire drill) in 2020?

- Yes (1)
- No (2)
- Don't know (3)

Display This Question:

If Was the plan tested in practice (like you would practice a fire drill) in 2020? = Yes

How many times was the plan tested? _______________________________

Did you ever modify your original plan in 2020?

- Yes (1)
- No (2)
- Don't know (3)

Is there a plan for an internal surge workforce in the event that it is needed?

- Yes (1)
- No (2)
- Don't know (3)

Display This Question:

If Is there a plan for an internal surge workforce in the event that it is needed? = Yes

What % of staff did/does the RACF anticipate would need to be furloughed in the event of an outbreak (and need to be replaced)?

|  | Up to 30% (1) | 30% to 50% (2) | > 50% (3) |
| --- | --- | --- | --- |
| Before July 2020 (1) |  |  |  |
| After July 2020 (2) |  |  |  |

Display This Question:

If Is there a plan for an internal surge workforce in the event that it is needed? = Yes

Did the surge workforce staff receive specific training on their roles as part of a surge workforce?

- Yes (1)
- No (2)
- Don't know (3)

Does the RACF have a formal COVID-19 communications plan which details levels of communication e.g., to residents, families and staff?

- Yes (1)
- No (2)
- Don't know (3)

What is your overall level of confidence that the outbreak management plan is fit for purpose and that you can rely on it to guide you and the RACF through an outbreak?

Please drag the blue circle to the position on the scale of 1 (low level of confidence) to 10 (high level of confidence).

|  | **Low level of confidence** | | | | | | **High level of confidence** | | | | | |
| --- | --- | --- | --- | --- | --- | --- | --- | --- | --- | --- | --- | --- |
|  | | 1 | 2 | 3 | 4 | 5 | | 6 | 7 | 8 | 9 | 10 |

| Confidence | 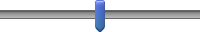 |
| --- | --- |

Lines of defence

In a recent review into major COVID-19 outbreaks in Australia, the following ***“lines of defence”*** were highlighted as critical to the effective and successful management of an outbreak. 
 
They are all critical. Based on your knowledge and experience, please rank them in order of their importance. For example, if you believe effective communication is most important, please rank as 1 or if you think it’s least important, please rank as 7.

**Rank all of the “lines of defence” for outbreak management from 1 (top) to 7 (bottom) by dragging each item to the desired position on the list – and add comments on any of them if you wish to in the box to the right.**

______ Leadership and management (1)

______ Effective communication (2)

______ Planning and preparation (3)

______ Infection control (4)

______ Pathology testing regime (5)

______ Workforce (6)

______ Emergency management (7)

Priorities for review

We are keen to understand other factors which you think are important for the review to focus on, although they may not be mentioned in the previous list.    Based on your experience with COVID-19 to date, **please provide your top 3 priorities to the review for its consideration.**

- Priority #1 (1) ________________________________________________
- Priority #2 (2) ________________________________________________
- Priority #3 (3) ________________________________________________

And in brief (just a few key words) **tell us why these priorities matter** to you and/or your RACF.

- Priority #1 (1) ________________________________________________
- Priority #2 (2) ________________________________________________
- Priority #3 (3) ________________________________________________

Guidance satisfaction

During the course of the COVID-19 pandemic, there has been much advice, direction and guidance to the sector from the Commonwealth Department of Health, State/Territory Health Departments, the Public Health Units and the Aged Care Quality and Safety Commission.    **Based on your experience, how would you rate your overall satisfaction with this information and the way it was delivered:**

|  | Very dissatisfied (1) | Quite dissatisfied (2) | Neutral (3) | Quite satisfied (4) | Very satisfied (5) | *Not aware of advice, direction or guidance* (6) |
| --- | --- | --- | --- | --- | --- | --- |
| **Commonwealth Department of Health** (1) |  |  |  |  |  |  |
| **State/Territory Department of Health** (2) |  |  |  |  |  |  |
| **Public Health Unit** (3) |  |  |  |  |  |  |
| **Aged Care Quality and Safety Commission** (4) |  |  |  |  |  |  |

Training

Did staff in your RACF complete **on-line training** in COVID-19 specific infection control during 2020?

- All (above 90%) (1)
- Most (61-90%) (2)
- Some (40-60%) (3)
- Few (under 40%) (4)

Did staff in your RACF receive **face to face training** in COVID-19 specific infection control during 2020?

- All (above 90%) (1)
- Most (61-90%) (2)
- Some (40-60%) (3)
- Few (under 40%) (4)

Outbreak

During 2020, did your RACF experience an outbreak (at least one confirmed resident or staff COVID-19 case)?

- Yes (1)
- No (2)
- Don't know (3)

Outbreak questions

How many confirmed **resident cases** did your RACF experience? ___________________________

How many confirmed **staff cases** did your RACF experience? ______________________________

How many residents died? __________________________________

Were you able to maintain an adequate workforce during the outbreak?

- All the time (above 90%) (1)
- Most of the time (61-90%) (2)
- Some of the time (40-60%) (3)
- Infrequently (under 40%) (4)

Did any of the RACF's permanent staff not attend for work as a result of concerns for their health and wellbeing?

- A few (under 20%) (1)
- A significant number (above 20%) (2)

Were staff able to maintain the required care documentation during the course of the outbreak?

- All the time (above 90%) (1)
- Most of the time (61-90%) (2)
- Some of the time (40-60%) (3)
- Infrequently (under 40%) (4)

Were you able to access adequate PPE supplies in a timely manner?

- All the time (above 90%) (1)
- Most of the time (61-90%) (2)
- Some of the time (40-60%) (3)
- Infrequently (under 40%) (4)

Does the RACF use an electronic or paper-based system for recording care delivery?

- Electronic system (1)
- Paper-based system (2)
- Some of both (3)

Did the residents wear a form of identification (e.g., wristband with name) during the course of the outbreak?

- Yes (1)
- No (2)
- Don't know (3)

Did the RACF engage additional staff to help reduce loneliness for residents (e.g., providing additional activities, engaging with residents, assisting with resident to family communications)?

- Yes (1)
- No (2)
- Don't know (3)

Were processes in place to strictly monitor residents' daily intake of food and fluids?

- Yes (1)
- No (2)
- Don't know (3)

As you reflect on the outbreak, in hindsight, how would you now rate your level of preparedness to manage that outbreak at that time?
Please drag the blue circle to the position on the scale of 1 (low preparedness) to 10 (high preparedness).

|  | | **Low** | | | | | | **High** | | | | |
| --- | --- | --- | --- | --- | --- | --- | --- | --- | --- | --- | --- | --- |
|  | 1 | | 2 | 3 | 4 | 5 | 6 | | 7 | 8 | 9 | 10 |

| Preparedness | 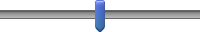 |
| --- | --- |

If there were 3 things that you could change, in the event of a future outbreak, what would they be?

- 1. (1) ________________________________________________
- 2. (2) ________________________________________________
- 3. (3) ________________________________________________

How would you rate support from **local area health services?**
Please drag the blue circle to the position on the scale of 1 (poor) to 5 (excellent) or indicate if not applicable.

|  | Poor | | | Satisfactory | | Excellent | | Not Applicable | |
| --- | --- | --- | --- | --- | --- | --- | --- | --- | --- |
|  | | 1 | 2 | | 3 | | 4 | | 5 |

| Support | 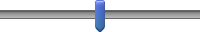 |
| --- | --- |

How would you rate support from **in-reach services?**
Please drag the blue circle to the position on the scale of 1 (poor) to 5 (excellent) or indicate if not applicable.

|  | Poor | | | Satisfactory | | Excellent | | Not Applicable | |
| --- | --- | --- | --- | --- | --- | --- | --- | --- | --- |
|  | | 1 | 2 | | 3 | | 4 | | 5 |

| Support | 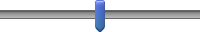 |
| --- | --- |

Following the outbreak, did your RACF undertake a structured review of the way in which the outbreak had been managed to understand what worked well and what needed to be improved?

- Yes (1)
- No (2)
- Don't know (3)

Display This Question:

If Following the outbreak, did your RACF undertake a structured review of the way in which the outbr... = Yes

Was the report provided to the organisation's Executive Committee?

- Yes (1)
- No (2)
- Don't know (3)

Display This Question:

If Following the outbreak, did your RACF undertake a structured review of the way in which the outbr... = Yes

Was the report provided to the Board (or one of its Committees)?

- Yes (1)
- No (2)
- Don't know (3)

Comments

Do you have any suggestions or comments you would like to share to help improve management of infectious disease outbreaks in the future?

________________________________________________________________

________________________________________________________________

________________________________________________________________

Would you be willing to be contacted about future research projects on this topic?
(This does not obligate you to participate in any future research)

- Yes (1)
- No (2)

Display This Question:

If Would you be willing to be contacted about future research projects on this topic? (This does not... = Yes

As you indicated you are willing to be contacted about future research, please provide your email address: ______________________________________________

Ending

Thank you for completing the survey. Your feedback is invaluable.

We appreciate your time and commitment to helping improve system responses to potential COVID-19 outbreaks in residential care.

Kind regards,

**The Independent Reviewers**

**SUPPLEMENTARY TABLE 1.**

Exploratory analysis^1^ of associations between nine organisational variables, in Australian RACFs included in the survey, (n=331), and COVID-19 outbreaks (official definition)^2^.

| **Variable**^3^ | | **Adjusted  Relative Risk (aRR)** | **95% confidence interval** | **p-value** |
| --- | --- | --- | --- | --- |
| *RACF Location* | Victoria | 12.08 | 4.74, 30.75 | <.001 |
|  | Not Victoria | 1.00 (ref) |  |  |
| *Rurality of RACF* | Regional/Rural/Remote | 0.52 | 0.23, 1.17 | .11 |
|  | Metropolitan | 1.00 (ref) |  |  |
| *RACF Ownership* | For-profit | 1.75 | 0.79, 3.92 | .17 |
|  | Not-for-profit/public | 1.00 (ref) |  |  |
| *RACF organisation* | Standalone | 0.86 | 0.44, 1.66 | .65 |
|  | Part of a group | 1.00 (ref) |  |  |
| *Outbreaks management plan (OMP)* | |  |  | .59 |
|  | None | 1.20 | 0.58, 2.49 | .62 |
|  | Late | 1.44 | 0.70, 2.94 | .32 |
|  | Early | 1.00 (ref) |  |  |
| *OMP practised* | No plan/not practised | 0.98 | 0.46, 2.06 | .95 |
|  | Tested | 1.00 (ref) |  |  |
| *Registered places* | >=75 | 1.00 | 0.54, 1.83 | .99 |
|  | <75 | 1.00 (ref) |  |  |
| *Shared facilities* | |  |  | .53 |
|  | Bedroom & bathroom | 1.24 | 0.68, 2.26 | .48 |
|  | Either | 0.48 | 0.07, 3.39 | .46 |
|  | None | 1.00 (ref) |  |  |
| *Staff IPC training: all Australian RACFs n=331; with outbreaks: n=32* | | | |  |
| *On-line* | Less than 90% | 0.96 | 0.50, 1.85 | .90 |
|  | 90% or more | 1.00 (ref) |  |  |
| *Face-to-face* | Less than 90% | 2.10 | 1.17, 3.79 | .013 |
|  | 90% or more | 1.00 (ref) |  |  |
| *Staff IPC training: Victorian RACFs n=91; with outbreaks: n=27^4^* | | |  |  |
| *On-line* | Less than 90% | 0.75 | 0.38, 1.47 | .40 |
|  | 90% or more | 1.00 (ref) |  |  |
| *Face-to-face* | Less than 90% | 2.20 | 1.15, 4.19 | .017 |
|  | 90% or more | 1.00 (ref) |  |  |

*Notes:*

1. Exploratory analyses were designed to identify potential organisational variables associated with outbreaks, in all RACFs included in the survey (n-331), of which 32 experienced one or more COVID-19 cases. The nine variables were included in a generalised linear model (modified Poisson approach, with a log-link function and robust standard errors to estimate relative risks and corresponding 95% confidence intervals).
2. The official definition of a COVID-19 outbreak was at least one case in a staff member, resident or frequent visitor. Outbreaks occurred in 32 of the 331 RACFs included in this survey.
3. See text for description of variables.
4. A separate exploratory analysis was performed based on organisational factors in Victorian RACFs (n=91), of which 27 experienced COVID-19 outbreaks. As for the analysis of all Australian RACFs, no significant correlations were identified between outbreaks and any variables in Victorian RACFs, other than staff face-to-face IPC training.

**SUPPLEMENTARY TABLE 2.**

Exploratory analysis^1^ of associations between nine organisational variables, in Australian RACFs included in the survey, (n=331), and COVID-19 outbreaks (restricted definition: five cases or more)^2^.

| **Variable**^3^ | | **Adjusted  Relative Risk (aRR)** | **95% confidence interval** | **p-value^3^** |
| --- | --- | --- | --- | --- |
| *RACF Location* | Victoria | 35.95 | 4.25, 304.12 | .001 |
|  | Not Victoria | 1.00 (ref) |  |  |
| *Rurality of RACF* | Regional/Rural/Remote | 0.05 | 0.01, 0.32 | .002 |
|  | Metropolitan | 1.00 (ref) |  |  |
| *RACF Ownership* | For-profit | 1.51 | 0.57, 3.97 | .40 |
|  | Not-for-profit/public | 1.00 (ref) |  |  |
| *RACF organisation* | Standalone | 1.38 | 0.54, 3.53 | .50 |
|  | Part of a group | 1.00 (ref) |  |  |
| *Outbreak management plan (OMP)* | |  |  | .64 |
|  | None | 0.80 | 0.33, 1.93 | .61 |
|  | Late | 1.27 | 0.40, 3.97 | .68 |
|  | Early | 1.00 (ref) |  |  |
| *OMP practised* | No plan/not practised | 1.32 | 0.44, 3.99 | .62 |
|  | Tested | 1.00 (ref) |  |  |
| *Registered places* | >=75 | 0.69 | 0.26, 1.81 | .45 |
|  | <75 | 1.00 (ref) |  |  |
| *Shared facilities* | |  |  | .98 |
|  | Bedroom & bathroom | 1.06 | 0.50, 2.26 | .87 |
|  | Either | 0.91 | 0.14, 6.12 | .93 |
|  | None | 1.00 (ref) |  |  |
| *Staff IPC training: all Australian RACFs n=331; with outbreaks: n=32* | | | |  |
| *On-line* | Less than 90% | 1.51 | 0.58, 3.92 | .42 |
|  | 90% or more | 1.00 (ref) |  |  |
| *Face-to-face* | Less than 90% | 2.58 | 1.00, 6.67 | .051 |
|  | 90% or more | 1.00 (ref) |  |  |

*Notes:*

1. See Suppl Table 1 for description of exploratory analyses and variables included in the model.
2. Nineteen RACFs experienced COVID-19 outbreaks of five or more cases in a staff member, resident, or frequent visitor. The definition assumed that this number of cases reflected COVID-19 transmission within the RACF.
3. *The number of outcomes of interest is small (n=19), the definition of an outbreak arbitrary, and the timing of cases relative to outbreaks unknown, so the results should be interpreted with caution.*
